# mRNA and Protein Subunit COVID-19 Vaccine Reactogenicity and the Relationship to Productivity for Healthcare Workers and First Responders

**DOI:** 10.1101/2025.08.25.25334392

**Authors:** Sarang K. Yoon, Andrew L. Phillips, Sarah Calhoun, Dawn Odom, Ryan Ziemiecki, Laurin Jackson, Jacob Mckell, Joshua Griffin, Ji Min Choi, Barbara A. Prillaman, Riley Campbell, Beth Hahn, Brandy Warren, Jonathan Fix, Matthew D. Rousculp, German L. Ellsworth

## Abstract

**Objectives:** To compare the percentage of participants that experienced any solicited systemic reactogenicity symptom after one dose of the updated 2024–2025 adjuvanted protein-based Novavax (NVX; JN.1) or Pfizer–BioNTech mRNA (PFZ; KP.2) COVID-19 vaccine within 2- and 7-days post-vaccination and examine its disrupting impact on daily life.

**Methods:** In this prospective, interventional, real-world study (SHIELD/NCT06633835), previously vaccinated healthcare workers (HCWs) and first responders (FRs) from Salt Lake City, Utah, USA, and surrounding areas self-selected to receive NVX or PFZ. At 2- and 7-days post-vaccination, participants self-reported on 11 vaccine-related reactogenicity symptoms and functional impairment using a 5-item modified Sheehan Disability Scale.

**Results:** 588 participants completed the Day 2 questionnaire (NVX, n=219; PFZ, n=369); 583 completed the Day 7 questionnaire (NVX, n=217; PFZ, n=366). Two-days post-vaccination, NVX recipients were less likely to report a systemic (OR=0.40; 95% CI, 0.21–0.75) or local (OR=0.10; 95% CI, 0.30–0.33) reactogenicity symptom and lost 50% fewer work hours (0.7 vs 1.4 h) and 66% fewer productive hours (0.8 vs 2.4 h) than PFZ recipients. There was little to no difference between vaccines at 7-days post-vaccination.

**Conclusion:** Fewer reactogenicity symptoms following COVID-19 vaccination suggest use of the NVX vaccine as an immunization option with a lower disruption to daily life.

## Introduction

The global COVID-19 pandemic led to the unprecedented development and deployment of multiple vaccines beginning in late 2020.^1,2^ These vaccines, based on diverse platforms including messenger ribonucleic acid (mRNA), protein subunit, and recombinant vectors, have proven effective in preventing severe illness and reducing transmission.^2–4^ Alongside their demonstrated efficacy, there is an increased emphasis on understanding vaccine safety and tolerability.^5^ Reactogenicity is a key component of postmarketing surveillance, particularly as new vaccine formulations and platforms continue to be introduced.^5^

The Centers for Disease Control and Prevention (CDC) recommended and approved three COVID-19 vaccines for the 2024–2025 season: the Pfizer–BioNTech mRNA vaccine (PFZ), the Moderna mRNA vaccine, and the Novavax protein-based vaccine (NVX).^6^ Both vaccine types have shown ≥90% efficacy to protect against severe COVID-19.^3,4^

Recipients of all three vaccine platforms have experienced transient reactogenicity events that are most prevalent within 2–3 days following initial and subsequent COVID-19 vaccinations^3,5,10–16^ and may increase with additional doses.^11^ Reactogenicity symptoms include local/injection-site reactions (e.g., pain, tenderness, swelling, or erythema) or systemic reactions (e.g., fever, headache, fatigue, malaise, muscle pain, joint pain, or nausea/vomiting).^3,5,10–16^ In a general population study, participants who received a protein-based vaccine reported lower reactogenicity and fewer impacts on their daily activities than participants who received an mRNA-based vaccine.^17,18^ Evidence also suggests that reactogenicity is lower in people who received additional protein-based vaccines than mRNA-based vaccines.^19,20^

Studies have shown that these reactogenicity symptoms can affect the ability of healthcare workers (HCWs) and first responders (FRs) to perform their duties.^16,21^ We designed the SHIELD (**S**tudy of **H**ealthcare Workers and First Responders **I**nvestigating **E**ffects Systemic and **L**ocal of COVID-19 Vaccine **D**oses) study to examine the impact of vaccine reactogenicity on productivity among US HCWs and FRs. To address this question, we examined the difference in percentage of participants that experienced any solicited systemic reactogenicity symptom (i.e., fever, fatigue, malaise, muscle pain, joint pain, nausea/vomiting, and headache) after receiving a single dose of an updated 2024–2025 NVX or PFZ within 2- and 7-days post-vaccination.

## Methods

### Study design and participants

We conducted a prospective, interventional, real-world study at the University of Utah between September 20, 2024, and December 17, 2024. We recruited HCWs and FRs from Salt Lake City, Utah, United States, and surrounding areas. HCWs and FRs were defined as anyone having direct face-to-face contact (defined as being within 3 feet, or about an arm’s length) with patients as part of their full- or part-time (≥20 h per week) job responsibilities. Figure S1 depicts the study design.

In addition to being an HCW/FR, inclusion criteria included age of ≥18 years; previous vaccination with at least one COVID-19 vaccine dose in the past 4 years; ability and willingness to comply with all study requirements; ability to understand and provide informed consent; and access to a smartphone, tablet, or computer and being comfortable receiving text messages/emails. Exclusion criteria included receipt of a COVID-19 vaccine within 60 days of enrollment; participation in another vaccine or investigational product trial within 90 days of study enrollment; history of self-reported severe allergic reaction to a COVID-19 vaccine; and plans to receive additional vaccines within 7-days post study vaccination (co-administration on vaccination day was permissible).

Participants self-selected their preferred COVID-19 vaccine of either NVX (2024-2025 JN.1 formulation) or PFZ (KP.2 formulation) up to the time of vaccine administration. Publicly available vaccine information was provided to inform the participant about the different vaccine platforms.^22,23^ Choice of vaccine was made solely by the participant. Investigators abstained from offering recommendations or influencing vaccine choice to maintain study objectivity. Eligible participants self-scheduled their own study visit appointment online. At the study visit, study personnel obtained informed consent in person and queried the state immunization registry to confirm COVID-19 vaccination history (i.e., received at least one dose of any COVID-19 vaccine and most recent dose was >60 days of enrollment). After receiving consent and determining eligibility, a licensed, study-affiliated medical provider verified the chosen vaccine and administered a single dose of the 0.5 mL NVX or 0.3 mL PFZ intramuscularly into the deltoid. After vaccination, participants were observed for 15 min to monitor for adverse reactions (i.e., syncope and allergic reactions). During this observation period, study personnel educated participants on adverse events such as myocarditis and pericarditis, upcoming study activities, and how to contact the study team. Additionally, participants were advised to contact the study physicians for medical advice if they experienced adverse reactions.

### Ethics

The study protocol, including participant compensation, was approved by the institutional review board (IRB) at the University of Utah (IRB #00181626; ClinicalTrials.gov identifier: NCT06633835).

### Reactogenicity assessments

Two days following vaccination, participants received an electronic questionnaire via text message and/or email. If not completed, participants received daily reminders to complete the questionnaire until the survey closed at midnight 2-days after it was first sent. The questionnaire collected sociodemographic information, daily activities/work, medical history, occupational history, and post-vaccination symptoms, which included 11 vaccine-related local or systemic reactogenicity symptoms (Figure S1). The questionnaire used a graded response scale from 0 (symptom not present) to 3 (significant symptom or impairment due to the symptom). Participants were compensated with a generic electronic gift card ($60) for the completion of each questionnaire (e.g., one for Day 2 and one for Day 7). At day 7 after vaccination, participants received a follow-up message asking about reactogenicity and functional impairment from days 3 to 7, using the same questions as the Day 2 survey.

Functional impairment was captured using the Sheehan Disability Scale (SDS; modified), a 5- item questionnaire consisting of three domains measuring disability and two questions measuring presenteeism and absenteeism.^24,25^ For each domain (work/school, social life, and family life/home responsibilities), the SDS^24,25^ assessed the degree to which vaccine symptoms caused disruption using a 0-to-10-point scale, with a higher score indicating greater functional disability.^26^ A total score was calculated by adding each domain.

In addition to the reactogenicity and functional impairment questions, self-reported information on sociodemographic characteristics, medical history (including past COVID-19 and influenza history), and employment status were collected on Day 0 and Day 2.

### Objectives

The primary objective was to determine the difference in percentage of participants who experienced any solicited systemic reactogenicity symptoms after receiving either a single dose of NVX or PFZ within 2-days post-vaccination. Secondary objectives included evaluating the following: percentage of participants who experienced any local reactogenicity symptoms, participant’s mean number of any systemic reactogenicity symptoms, percentage of participants reporting systemic or local reactogenicity of grade 2 or higher, and disruptions in participants’ work/social/family life, all within 2-days post-vaccination.

### Statistical analysis

The sample size of this study was determined to ensure power for the primary objective, with assumptions based on a previous COVID-19 vaccine reactogenicity observational study.^18^ We assumed that participants receiving PFZ would enroll faster than NVX participants and used a 2:1 ratio (PFZ:NVX) for enrollment. The minimum sample size required to assess a 15% absolute difference in the primary objective at 90% power using a 2-sided test with a 0.05 significance level was approximately 525 participants (PFZ: 350; NVX: 175). An assumed 20% loss in total participants was included in the estimation of the final target sample size of 440 PFZ and 220 NVX participants. Actual sample size, including the ratio of PFZ to NVX, was monitored closely during the study, and enrollment was stopped a few weeks early due to quicker enrollment and lower loss to follow-up than anticipated. Descriptive analyses were conducted for all questions. Comparative analyses were conducted on key study endpoints. Odds ratios (ORs), percentages, least square means, confidence intervals (CIs), and p values for the difference between vaccine groups were derived from logistic regression, negative binomial, or analysis of variance models, as appropriate, adjusted for age, sex, and whether the participant primarily worked in a hospital setting. Participants with incomplete responses were removed from analysis.

### Role of the funding source

Funding was provided by Novavax, Inc.

## Results

### Participant demographics

Of the 660 individuals screened, 588 enrolled participants received either NVX or PFZ. Within 15 min after vaccination, there were no adverse reactions reported. All 588 participants completed the Day 2 post-vaccination questionnaire (NVX, n=219; PFZ, n=369) and 583 (99%) went on to complete the Day 7 post-vaccination questionnaire (NVX, n=217; PFZ, n=366). Co-administration of an influenza vaccine was low among participants (NVX, 0.46%; PFZ, 0.54%). Figure 1 shows the participant screening and inclusion process.

**Figure 1.**
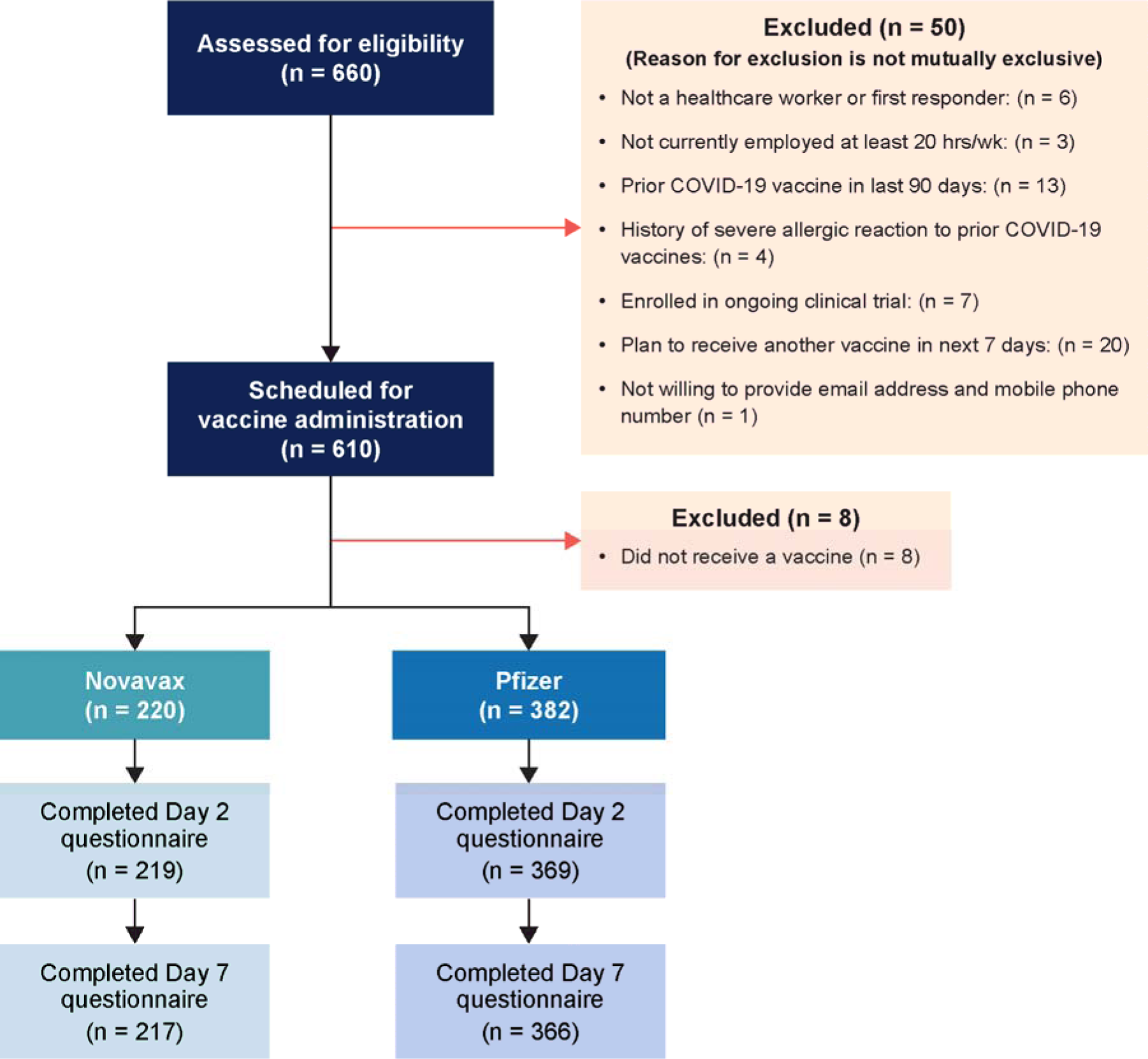
Participant screening and inclusion. NVX = Novavax protein subunit COVID-19 vaccine; PFZ = Pfizer-BioNTech mRNA vaccine.

Full participant demographics, medical history, and occupational history are reported in Table 1. Participants’ mean age (standard deviation) was 36.4 (11.0) years, and 70.1% identified as female. Participants were predominantly White (84.0%), and 11.6% identified as Hispanic. Within the sample, 82.8% worked in a hospital; 20.4% were physicians, 19.2% were nurses/nurse practitioners, and 11.9% were medical assistants. Baseline characteristics (e.g., age, gender, income) were balanced across the two vaccine populations, except for employment location and hospital setting. Although initial plans included propensity score analysis to adjust for baseline differences, the observed balance between groups supported the use of direct covariate adjustment as the more appropriate analytical approach.

**Table 1.** Participant demographics, medical history, and occupational history.

|  | **NVX (n=219)** | **PFZ (n=369)** | **Overall (n=588)** |
| --- | --- | --- | --- |
| **General demographics** |  |  |  |
| Mean age (SD) | 36.6 (10.65) | 36.3 (11.23) | 36.4 (11.01) |
| Gender identity, n (%) |  |  |  |
| Female | 150 (68.5) | 262 (71.0) | 412 (70.1) |
| Gender fluid | 0 (0.0) | 1 (0.3) | 1 (0.2) |
| Nonbinary | 5 (2.3) | 3 (0.8) | 8 (1.4) |
| Male | 63 (28.8) | 103 (27.9) | 166 (28.2) |
| Prefer not to answer | 1 (0.5) | 0 (0.0) | 1 (0.2) |
| Identifies as Hispanic | 25 (11.4) | 43 (11.7) | 68 (11.6) |
| Race |  |  |  |
| African American or Black | 1 (0.5) | 3 (0.8) | 4 (0.7) |
| Alaska Native, American Indian, or Native American | 4 (1.8) | 7 (1.9) | 11 (1.9) |
| Asian or Asian American | 21 (9.6) | 46 (12.5) | 67 (11.4) |
| Middle Eastern and/or North African | 3 (1.4) | 6 (1.6) | 9 (1.5) |
| Native Hawaiian and/or Pacific Islander | 1 (0.5) | 0 (0.0) | 1 (0.2) |
| White | 185 (84.5) | 309 (83.7) | 494 (84.0) |
| A race not listed | 8 (3.7) | 11 (3.0) | 19 (3.2) |
| I prefer not to answer | 4 (1.8) | 6 (1.6) | 10 (1.7) |
| Highest educational level, n (%) |  |  |  |
| Less than high school | 1 (0.5) | 0 (0.0) | 1 (0.2) |
| High school diploma or equivalent (for example, GED) | 6 (2.7) | 5 (1.4) | 11 (1.9) |
| Some college | 24 (11.0) | 43 (11.7) | 67 (11.4) |
| College degree | 67 (30.6) | 113 (30.6) | 180 (30.6) |
| Professional or advanced degree | 121 (55.3) | 208 (56.4) | 329 (56.0) |
| Marital status, n (%) |  |  |  |
| Single/never married | 79 (36.1) | 126 (34.1) | 205 (34.9) |
| Married/living as married/civil partnership | 120 (54.8) | 223 (60.4) | 343 (58.3) |
| Divorced or separated | 17 (7.8) | 17 (4.6) | 34 (5.8) |
| Widowed/surviving partner | 0 (0.0) | 3 (0.8) | 3 (0.5) |
| Other | 2 (0.9) | 0 (0.0) | 2 (0.3) |
| Prefer not to answer | 1 (0.5) | 0 (0.0) | 1 (0.2) |
| Household size |  |  |  |
| 1 person (self) | 37 (16.9) | 60 (16.3) | 97 (16.5) |
| 2 people | 78 (35.6) | 136 (36.9) | 214 (36.4) |
| 3 people | 36 (16.4) | 63 (17.1) | 99 (16.8) |
| 4 people | 36 (16.4) | 63 (17.1) | 99 (16.8) |
| 5 people | 16 (7.3) | 27 (7.3) | 43 (7.3) |
| >5 people | 16 (7.3) | 20 (5.4) | 36 (6.1) |
| Annual pre-tax income, n (%) |  |  |  |
| \$1 to \$9,999 | 4 (1.8) | 4 (1.1) | 8 (1.4) |
| \$10,000 to \$24,999 | 9 (4.1) | 12 (3.3) | 21 (3.6) |
| \$25,000 to \$49,999 | 32 (14.6) | 48 (13.0) | 80 (13.6) |
| \$50,000 to \$99,999 | 77 (35.2) | 148 (40.1) | 225 (38.3) |
| \$100,000 to \$199,999 | 54 (24.7) | 97 (26.3) | 151 (25.7) |
| \$200,000 to \$499,999 | 34 (15.5) | 38 (10.3) | 72 (12.2) |
| \$500,000 or more | 1 (0.5) | 4 (1.1) | 5 (0.9) |
| Prefer not to answer | 8 (3.7) | 18 (4.9) | 26 (4.4) |
| Community size, n (%) |  |  |  |
| Urban [a city or metropolitan area with 50,000 or more people] | 144 (65.8) | 249 (67.5) | 393 (66.8) |
| Suburban [a large residential area that surrounds metropolitan areas with at least 2,500 people but fewer than 50,000 residents] | 66 (30.1) | 113 (30.6) | 179 (30.4) |
| Rural [areas outside city and towns typically with open spaces, farms, forests, and natural scenery with limited infrastructure] | 8 (3.7) | 5 (1.4) | 13 (2.2) |
| Do not know | 1 (0.5) | 2 (0.5) | 3 (0.5) |
| Prefer not to answer | 0 (0.0) | 0 (0.0) | 0 (0.0) |
| **Medical history** |  |  |  |
| Type(s) of health insurance<sup>a</sup> n (%) |  |  |  |
| Private/commercial health insurance (individual or employer-provided insurance) | 208 (95.0) | 351 (95.1) | 559 (95.1) |
| Medicare or Medi-Gap | 0 (0.0) | 3 (0.8) | 3 (0.5) |
| Medicaid | 6 (2.7) | 6 (1.6) | 12 (2.0) |
| Military healthcare (TRICARE, VA, CHAMP-VA) | 2 (0.9) | 3 (0.8) | 5 (0.9) |
| Other coverage not listed | 1 (0.5) | 3 (0.8) | 4 (0.7) |
| No coverage of any type | 2 (0.9) | 4 (1.1) | 6 (1.0) |
| I prefer not to answer | 0 (0.0) | 1 (0.3) | 1 (0.2) |
| Medical conditions<sup>a</sup>, n (%) |  |  |  |
| Asthma | 32 (14.6) | 41 (11.1) | 73 (12.4) |
| Autoimmune diseases (such as lupus, rheumatic diseases) | 8 (3.7) | 16 (4.3) | 24 (4.1) |
| Active cancer (not in remission) | 0 (0.0) | 0 (0.0) | 0 (0.0) |
| Chronic kidney disease | 1 (0.5) | 0 (0.0) | 1 (0.2) |
| Chronic liver disease (such as chronic hepatitis) | 0 (0.0) | 2 (0.5) | 2 (0.3) |
| Chronic heart failure | 0 (0.0) | 0 (0.0) | 0 (0.0) |
| Central nervous system disorders (such as chronic neurological or neuromuscular disorders, dementia, or cerebrovascular disorders, epilepsy) | 0 (0.0) | 1 (0.3) | 1 (0.2) |
| Diabetes or other metabolic diseases | 4 (1.8) | 10 (2.7) | 14 (2.4) |
| HIV | 0 (0.0) | 0 (0.0) | 0 (0.0) |
| Hypertension (high blood pressure) | 19 (8.7) | 25 (6.8) | 44 (7.5) |
| Heart disease or condition (such as atherosclerosis, heart attack, stroke, congenital heart disease, coronary heart disease) | 1 (0.5) | 0 (0.0) | 1 (0.2) |
| Mental health disorders (such as psychiatric disorders, anxiety, depression) | 64 (29.2) | 85 (23.0) | 149 (25.3) |
| Obesity | 15 (6.8) | 35 (9.5) | 50 (8.5) |
| Organ transplantation | 0 (0.0) | 0 (0.0) | 0 (0.0) |
| Respiratory conditions (for example COPD, emphysema) | 0 (0.0) | 0 (0.0) | 0 (0.0) |
| Other congenital or acquired immunodeficiency or immunosuppression | 0 (0.0) | 1 (0.3) | 1 (0.2) |
| Other | 13 (5.9) | 18 (4.9) | 31 (5.3) |
| Prefer not to answer | 0 (0.0) | 1 (0.3) | 1 (0.2) |
| None | 113 (51.6) | 204 (55.3) | 317 (53.9) |
| **Occupational history** |  |  |  |
| Current role<sup>a</sup>, n (%) |  |  |  |
| Dentist | 0 (0.0) | 4 (1.1) | 4 (0.7) |
| Dental hygienist or assistant | 0 (0.0) | 2 (0.5) | 2 (0.3) |
| Dietician or nutritionist | 2 (0.9) | 7 (1.9) | 9 (1.5) |
| Fire service | 3 (1.4) | 3 (0.8) | 6 (1.0) |
| Medical assistant | 30 (13.7) | 40 (10.8) | 70 (11.9) |
| Non-fire emergency medical services (paramedic, EMT) | 8 (3.7) | 7 (1.9) | 15 (2.6) |
| Nurse (LPN) | 0 (0.0) | 0 (0.0) | 0 (0.0) |
| Nurse (RN) | 39 (17.8) | 57 (15.4) | 96 (16.3) |
| Nurse practitioner | 12 (5.5) | 5 (1.4) | 17 (2.9) |
| Occupational therapist | 6 (2.7) | 14 (3.8) | 20 (3.4) |
| Pharmacist or pharmacy staff | 8 (3.7) | 29 (7.9) | 37 (6.3) |
| Phlebotomist | 0 (0.0) | 1 (0.3) | 1 (0.2) |
| Physical therapist | 8 (3.7) | 31 (8.4) | 39 (6.6) |
| Physician | 51 (23.3) | 69 (18.7) | 120 (20.4) |
| Physician assistant | 12 (5.5) | 15 (4.1) | 27 (4.6) |
| Psychologist | 1 (0.5) | 5 (1.4) | 6 (1.0) |
| Radiology technician/ultrasound technician | 0 (0.0) | 0 (0.0) | 0 (0.0) |
| Receptionist/desk attendant/patient service representative | 3 (1.4) | 2 (0.5) | 5 (0.9) |
| Respiratory therapist | 2 (0.9) | 5 (1.4) | 7 (1.2) |
| Speech therapist | 5 (2.3) | 8 (2.2) | 13 (2.2) |
| Social worker | 2 (0.9) | 6 (1.6) | 8 (1.4) |
| Other | 35 (16.0) | 66 (17.9) | 101 (17.2) |
| Prefer not to answer | 0 (0.0) | 2 (0.5) | 2 (0.3) |
| Regularly works in a hospital n (%) | 172 (78.5) | 315 (85.4) | 487 (82.8) |
| Hospital work assignment, n (%) |  |  |  |
| Intensive care (e.g., ICU, NICU, PICU) | 46 (21.0) | 90 (24.4) | 136 (23.1) |
| Emergency department | 25 (11.4) | 39 (10.6) | 64 (10.9) |
| Maternity ward | 18 (8.2) | 23 (6.2) | 41 (7.0) |
| Medical/surgical ward | 63 (28.8) | 119 (32.2) | 182 (31.0) |
| Other | 76 (34.7) | 139 (37.7) | 215 (36.6) |
| Prefer not to answer | 0 (0.0) | 1 (0.3) | 1 (0.2) |
| Regularly works at a long-term care or other nursing facility, n (%) | 10 (4.6) | 20 (5.4) | 30 (5.1) |
| Regularly works in an outpatient care facility, n (%) | 121 (55.3) | 176 (47.7) | 297 (50.5) |
| Work location (outpatient)<sup>a</sup> n (%) |  |  |  |
| Doctor's office/clinic | 109 (49.8) | 154 (41.7) | 263 (44.7) |
| Urgent care clinic | 7 (3.2) | 4 (1.1) | 11 (1.9) |
| Other outpatient facility (e.g., dental, dialysis, surgical) | 36 (16.4) | 60 (16.3) | 96 (16.3) |
| None of these | 85 (38.8) | 167 (45.3) | 252 (42.9) |
| Prefer not to answer | 1 (0.5) | 1 (0.3) | 2 (0.3) |

COPD = chronic obstructive pulmonary disease; EMT = emergency medical technicians; GED = General Educational Development; ICU = intensive care unit; LPN = licensed practical nurse; NICU = neonatal intensive care unit; NVX = Novavax protein subunit vaccine; PFZ = Pfizer-BioNTech mRNA vaccine; PICU = pediatric intensive care unit; RN = registered nurse; SD = standard deviation; VA = Veterans Affairs.
<sup>a</sup>Participants were allowed to select all that applied; therefore, percentages may add up to over 100%.
Note: Population is of participants completing the Day 2 post-vaccination questionnaire.

### Systemic reactogenicity symptoms

Within 2-days post-vaccination, fewer NVX recipients (73.6%) reported experiencing at least one systemic reactogenicity symptom (i.e., grade ≥1) compared with PFZ recipients (87.5%) (absolute difference: 13.8%, p=0.0044) (Table S1 and Figure 2A). Participants vaccinated with NVX were significantly less likely to report systemic reactogenicity symptoms within 2-days compared with those who received PFZ (OR=0.40; 95% CI, 0.21–0.75).

**Figure 2.**
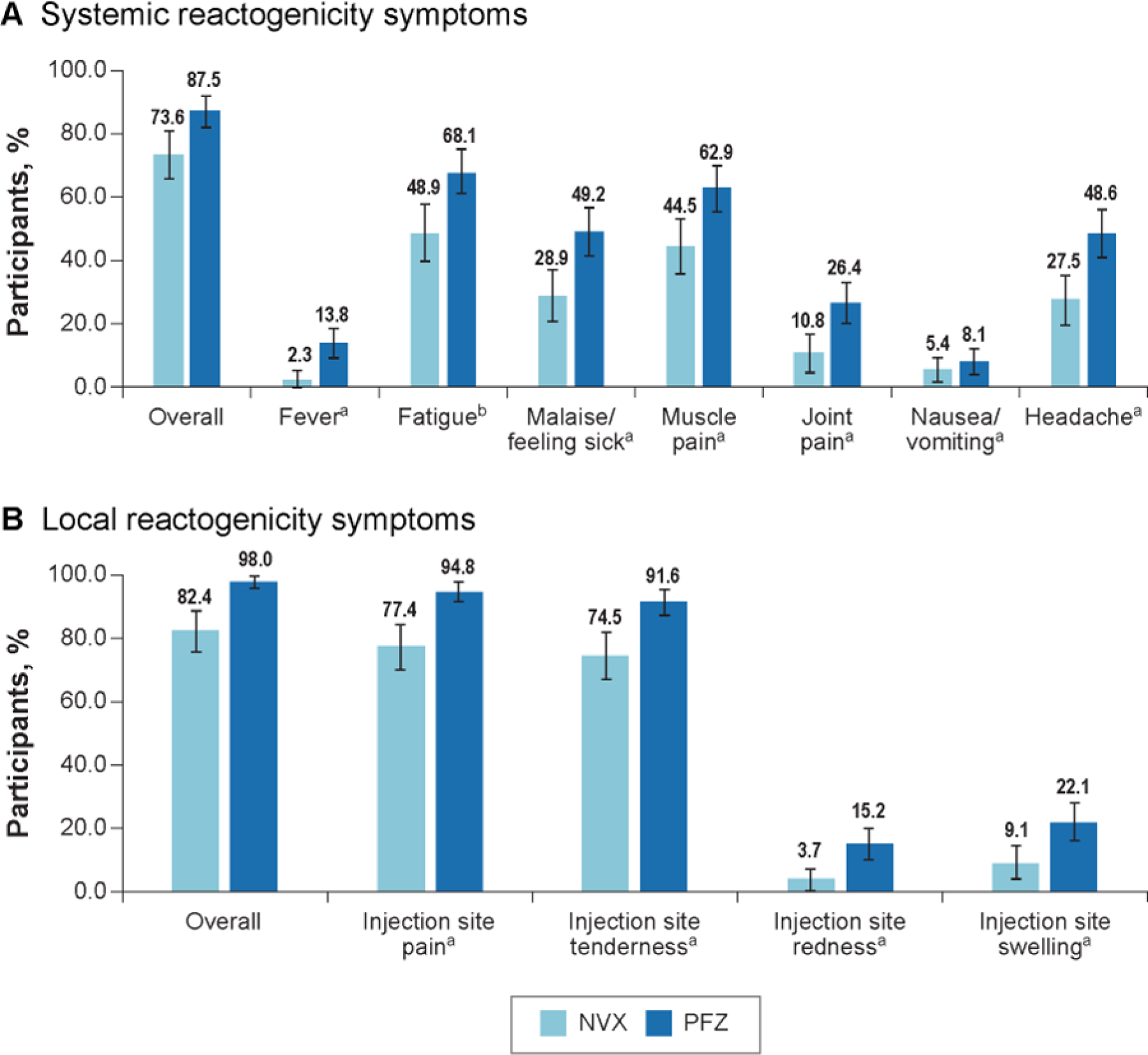
Percentage of participants with local and systemic reactogenicity symptoms 2-days post-vaccination. PFZ = Pfizer-BioNTech mRNA vaccine; NVX = Novavax protein-based COVID-19 vaccine. ^a^ Five participants were removed from the analysis sample due to incomplete responses (Novavax: 1 removed due to missing sex; Pfizer: 4 removed for missing hospital work setting). ^b^ Six participants were removed from the analysis sample due to incomplete responses (Novavax: 1 removed due to missing sex; Pfizer: 1 removed for incomplete reactogenicity, 4 removed for missing hospital work setting).

The mean number of systemic reactogenicity symptoms reported by participants who received NVX was 1.7 (95% CI, 1.5–2.0) compared with 2.8 (95% CI, 2.5–3.1) reported by participants who received PFZ (Table S2). After adjustment, participants who received NVX had, on average, 1.0 fewer systemic symptoms (95% CI, −1.4 to −0.6; p< 0.0001) than those who received PFZ.

After covariate adjustment, NVX recipients were less likely than PFZ participants to experience headache (OR=0.40; 95% CI, 0.24–0.67), malaise/feeling sick (OR=0.42; 95% CI, 0.25–0.70), fatigue (OR=0.45; 95% CI, 0.27–0.73), muscle pain (OR=0.47; 95% CI, 0.29–0.77, p=0.0024), and joint pain (OR=0.34; 95% CI, 0.16–0.71) (Figure 2A). Compared with recipients of PFZ, recipients of NVX also experienced less fever and nausea/vomiting (absolute difference: 11.5% and 2.6%, respectively).

Participants who received NVX as an additional dose had lower odds of experiencing a grade 2 or higher systemic reactogenicity symptom within 2-days post-vaccination than participants who received PFZ as an additional dose (OR=0.30; 95% CI, 0.17–0.52) (Table S3 and Figure 3).

**Figure 3.**
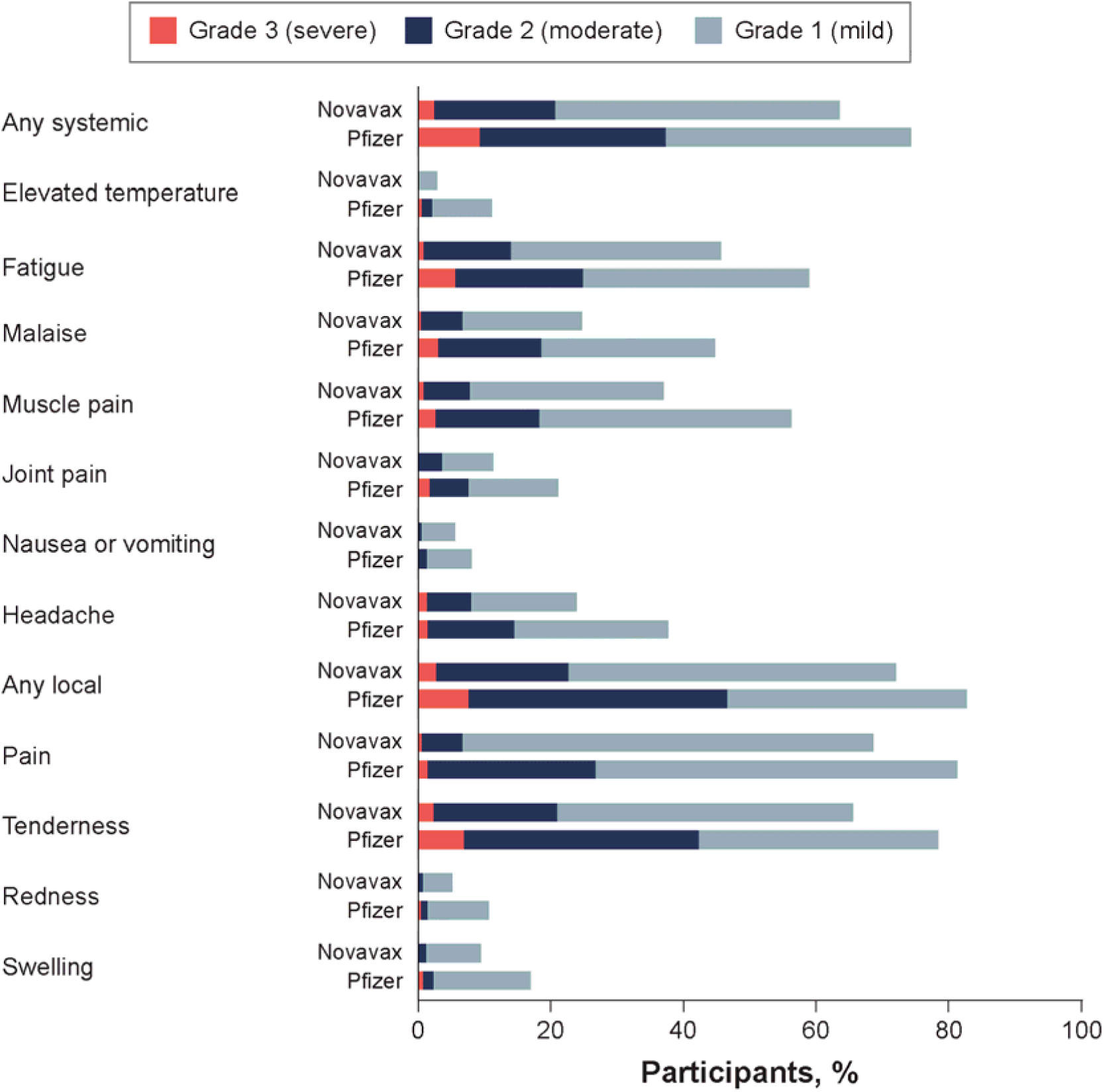
Systemic and local reactogenicity events after Novavax or Pfizer vaccines within 2-days post-vaccination.

Between 3- and 7-days post-vaccination, 36.4% of NVX recipients reported experiencing continued systemic symptoms compared with 42.1% of PFZ recipients (Table S4). Overall symptoms were transient, with <17% of all recipients reporting fever, malaise/feeling sick, muscle pain, nausea/vomiting, or joint pain and <29% of all previously symptomatic recipients reporting headache or fatigue on the Day 7 questionnaire. Among study participants who initially reported any of the solicited systemic symptoms on the Day 2 questionnaire, NVX vaccinees compared with PFZ vaccinees reported fewer headaches (10.1% vs 16.4%), muscle pain (10.6% vs 13.1%), malaise/feeling sick (9.7% vs 12.8%), fever (0.0% vs 1.1%), and fatigue (22.1% vs 26.0%) on days 3–7.

### Local reactogenicity symptoms

After covariate adjustment, 82.4% of NVX recipients and 98.0% of PFZ recipients reported experiencing at least one local reactogenicity symptom at day 2 post-vaccination (absolute difference: 15.6%, p<0.0001) (Table S1 and Figure 2B). NVX recipients were significantly less likely to report a local reactogenicity symptom than PFZ recipients (OR=0.10; 95% CI, 0.03– 0.33). NVX recipients were less likely to experience all measured local symptoms, including pain (OR=0.19; 95% CI, 0.09–0.41), tenderness (OR=0.27; 95% CI, 0.13–0.54), swelling (OR=0.35; 95% CI, 0.17–0.74), and redness (OR=0.22; 95% CI, 0.07–0.63) at the injection site.

Results revealed that 22.8% of NVX recipients experienced at least one local reactogenicity symptom of grade 2 or higher compared with 57.7% of PFZ recipients after adjustment (Table S3 and Figure 3) (absolute difference: 34.8%, p<0.0001; OR=0.22; 95% CI, 0.12–0.38). Among participants who experienced a grade 1 or worse symptom on day 2 after vaccination, 18.4% of NVX recipients reported experiencing continued systemic symptoms on the Day 7 questionnaire compared with 28.4% of PFZ recipients (Table S4). Symptoms were transient among all study participants who initially reported any solicited local symptoms on the Day 2 questionnaire, with lower frequencies among NVX compared with PFZ recipients for pain (15.2% vs 21.9%), tenderness (15.7% vs 24.0%), and swelling (3.7% vs 5.7%) at the injection site reported between 3- and 7-days post-vaccination. Frequency of redness was similar among NVX and PFZ groups (3.7% vs 3.8%).

### Disruption of work, social, and family life

Two days following vaccination, participants reported that they lost, on average, 50% fewer work hours (0.7 vs 1.4) and 66% fewer productive hours (0.8 vs 2.4) after receiving NVX compared with PFZ (Table S5). Participants receiving either vaccine described their work as being disrupted by vaccine symptoms mildly (17.9% vs 28.4%) and moderately (5.0% vs 10.6%). Compared with NVX recipients, a greater proportion of PFZ recipients reported a disruption to work or school activities (39.3% vs 54.3%), social or leisure activities (57.5% vs 72.6%), and family or home responsibilities (51.2% vs 71.2%) (Figure 4 and Table S5).

**Figure 4.**
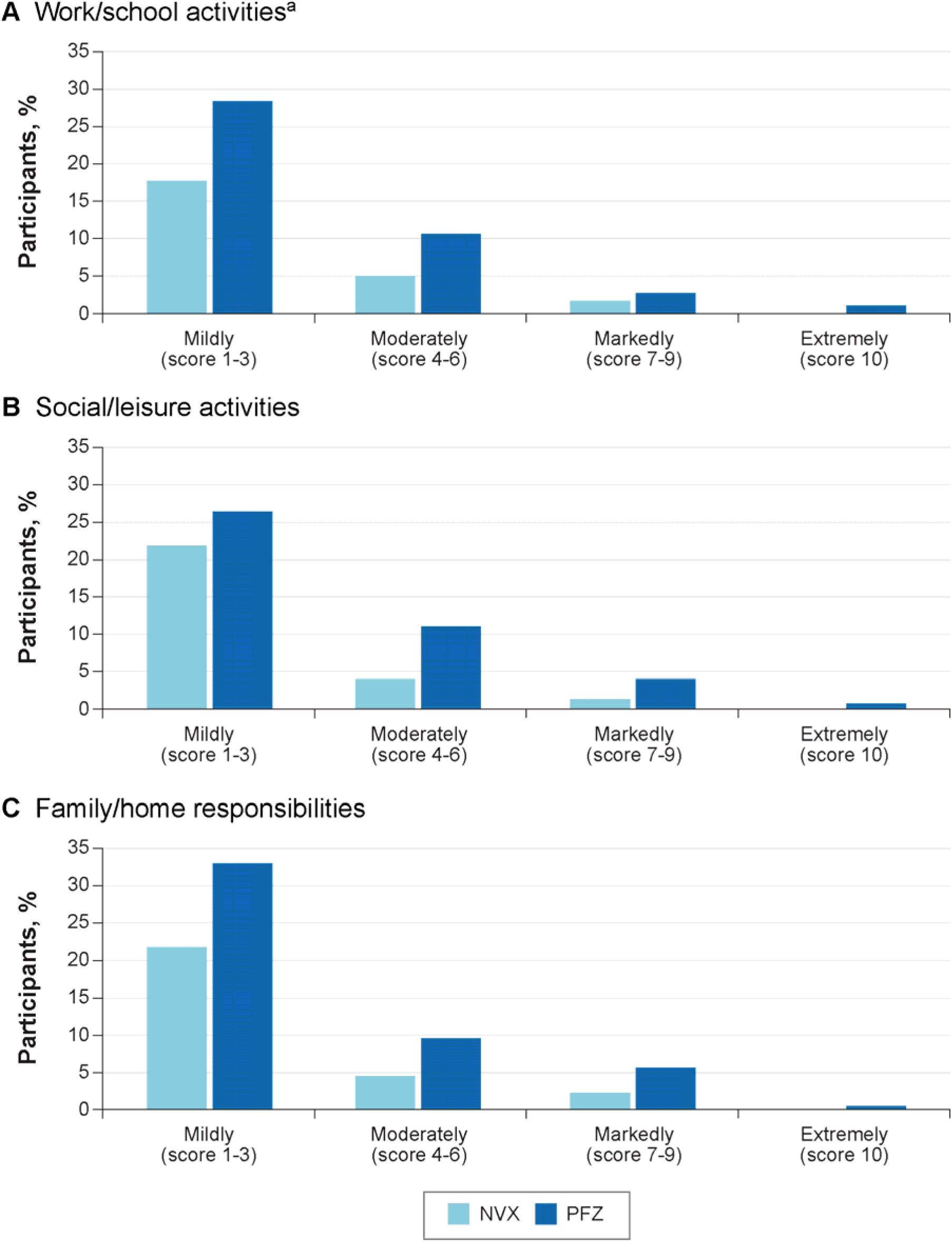
Disruption of activities caused by symptoms within the first 2-days post-vaccination. PFZ = Pfizer-BioNTech mRNA vaccine; NVX = Novavax protein-based COVID-19 vaccine. ^a^ 21.5% of NVX and 17.9% of PFZ recipients did not have work/school scheduled during the days following vaccination and were not included in these results.

The median total SDS score was lower among NVX recipients (0.0; interquartile range [IQR]=3.0]) than among PFZ recipients (2.0; IQR=7.0) (Table S5). After covariate adjustment, the difference in the least-squares mean was −2.6 (95% CI, −3.9 to −1.3, p=0.0001) for global impairment of work, social, and family life disruption at 2-days post-vaccination between vaccines and was significantly lower among participants who received NVX (1.9; 95% CI, 0.9– 2.9) than among those receiving PFZ (4.5; 95% CI, 3.7–5.4) (Table S6).

Descriptive analyses showed no meaningful differences at 3–7-days post-vaccination between the two vaccines in the average loss of work hours (1.1 [NVX] vs 0.8 [PFZ]) productive hours (1.1 [NVX] vs 1.0 [PFZ]), or median total SDS score (0.0, IQR=0.0 [NVX] vs 0.0, IQR=0.0 [PFZ]) (Table S7).

## Discussion

In this open-label, nonrandomized, interventional COVID-19 vaccine study of HCWs and FRs, NVX recipients were significantly less likely than PFZ recipients to experience systemic and local reactogenicity events within 2-days post-vaccination. This trend was consistent across most systemic and local symptoms, with the exception of nausea and vomiting. Regarding overall impact, NVX recipients reported a lower mean global impairment score related to work, social, and family life, with a statistically significant difference compared to PFZ recipients. For both vaccine groups, most reactogenicity events occurred within 2-days after vaccination and resolved by day 7.

The observed lower reactogenicity among NVX recipients compared with PFZ recipients may be attributed to the fundamental differences in vaccine platforms (i.e., utilizing lipid nanoparticles for PFZ vs Matrix-M saponin–based adjuvant for NVX; the spike protein production via mRNA genetic instructions vs premade proteins), kinetics of the immune response, and/or vaccine dosage (PFZ’s 30 μg in 0.3 mL vs NVX’s 5 μg spike protein + 50 μg Matrix-M in 0.5 mL).

Between 2020 and 2022, a systematic review and meta-analysis of blinded randomized controlled trials of globally available primary COVID-19 vaccines (viral vector, inactivated, mRNA, and protein subunit) versus placebo found that vaccine platform type significantly influences reactogenicity.^5^ A recent review examining the reactogenicity of a homologous or heterologous third doses of mRNA or NVX vaccines (following a primary series regimen) found that NVX was associated with the lowest frequency and severity of systemic and local reactogenicity events,^27^ in line with COV-BOOST phase 2 trial findings^20^ and real-world prospective studies.^28–30^ Interestingly, in our study, nausea/vomiting was not significantly different between vaccine groups—a pattern also observed in participants who received two doses of mRNA-1273 (Moderna) primary series and the NVX-CoV2373 as a third dose, as reported in the review by Marchese et al.^27^ While these differences are consistently observed, the underlying causes and mechanisms remain to be fully understood. Further comparative studies are needed to elucidate the underlying mechanisms between vaccine platforms, their respective immune responses, and resulting reactogenicity profiles.

Our findings add to the current literature by providing interventional evidence that supports and strengthens prior observational trends suggesting improved tolerability of NVX. While Rousculp et al.^17^ reported a trend toward lower impairment among NVX recipients in a prospective study, our study demonstrated a statistically significant reduction in global impairment scores 2-days post-vaccination (least-squares mean difference: –2.6; 95% CI, –3.9 to –1.3), highlighting that NVX recipients experienced fewer disruptions to daily functioning despite the presence of post-vaccination symptoms. Consideration of lower reactogenicity—in the context of effective vaccines—may be particularly relevant for high-risk occupations such as HCWs, where reduced post-vaccination impairment could increase acceptance among the workforce and institutions.

Reactogenicity may have implications for vaccine selection and uptake not only at the individual level but also among employers and institutions. While recent studies have confirmed that COVID-19 vaccines are safe and effective, some have also shown that side effects—though generally mild and transient—can affect the ability of working adults, including HCWs and FRs, to carry out their duties^21,31^; increase work absences^17,21,32,33^; and contribute to vaccine hesitancy.^34–36^ A 2021 worldwide scoping review found that up to 72% of HCWs expressed hesitancy about receiving a COVID-19 vaccine, with >75% of the studies citing concerns about safety, efficacy, and side effects as reasons for vaccine hesitancy; this finding mirrors reasons given from the general public for vaccine hesitancy, where 60% of individuals cited side effects, 48% cited safety, and 30% cited efficacy (“how well it works”).^37^ Another study in 2020 found that the most frequently reported side effects were fatigue (59%) and headache (52%), which although temporary, may reduce productivity and increase sick leave among healthcare personnel.^16^ Understanding and addressing post-vaccination reactogenicity symptoms is especially important in critical occupations such as healthcare and emergency response, where even short-term symptoms can have significant implications for workforce planning and staffing.

A key strength of the present study is its focus on vaccine impacts on a cohort of HCWs and FRs. Beyond showing significantly fewer reactogenic symptoms and lower burden of vaccination, this cohort is frequently seen as the most trusted group in providing vaccine guidance.^38,39^ Vaccine tolerability and concern of side effects are also leading reasons cited for vaccine hesitancy in the general public.^36,40^ Therefore, it is valuable for HCWs and FRs to be able to share their personal experiences and scientific evidence on the side effects of COVID-19 vaccines. Generalizability may be limited to these types of occupations or other high-risk occupation settings. Open-label and real-world studies offer valuable insights into how treatments perform outside of controlled settings. However, these study designs have inherent limitations. Open-label studies lack blinding, which can introduce performance and observer bias, reporting bias, and reduced internal validity. Real-world studies often lack randomization and are prone to selection bias, incomplete or inaccurate data, and heterogeneity in care and documentation—especially in studies relying on self-reported data.

This study was designed to address these limitations within the constraints of an interventional framework. For example, enrollment at the time of vaccination, combined with timely follow-up questionnaires (administered 2–7-days post-vaccination), aimed to reduce recall bias. To mitigate potential sources of bias, participants were allowed to select the vaccine of their choice, and study staff were not involved in the decision-making process. All participants were offered a choice among vaccines approved or authorized by the US Food and Drug Administration and recommended by the CDC’s Advisory Committee on Immunization Practices (ACIP). Additionally, all participants received identical surveys, were followed according to a consistent schedule, and were managed under standardized protocols across groups—measures intended to minimize performance and attrition bias. Overall, we observed exceptionally high compliance with study procedures and a very low attrition rate.

It is important to note that this study used a convenience sample, which may limit generalizability. Participants were recruited from a single geographic area (Salt Lake City, Utah, United States), and those who chose to participate may differ in important ways from those who declined. These findings may be specific to high-risk occupation settings, e.g., healthcare and emergency response, and may not be broadly applicable.

The results presented here build upon previously published studies that indicate the NVX vaccine has a lower reactogenicity and a lower impact on work absenteeism and productivity.^17,18^ This study is the first to evaluate the comparative side effects and impacts between the protein-based and mRNA vaccines in HCWs and FRs. Furthermore, these results offer valuable insights for guiding vaccine recommendations for both HCWs/FRs and healthcare administrators.

## Conclusion

The lower frequency and intensity of reactogenicity symptoms following COVID-19 vaccination observed in this open-label, real-world, interventional study suggests use of the adjuvanted NVX is a notable immunization option with lower reactogenicity symptoms and impacts on daily life; furthermore, it may be a valuable asset to address the current COVID-19 vaccine hesitancy that has led to immunization rates lower than influenza vaccination rates. These findings may provide meaningful insight for policymakers, clinicians, and hospital systems in future COVID-19 vaccination programs.

## Data Availability

Study information is available online at: https://www.clinicaltrials.gov/study/NCT06633835.
Requests submitted to the corresponding author will be considered upon publication of this article and de-identified participant data might be provided.

## Acknowledgements

The authors thank all the study participants who volunteered for this study. The authors would like to thank Justin Dash and Michael Remavich of RTI Health Solutions for their data management contributions. The authors would like to thank the SHIELD-Utah research staff and Bubba Brown from the Rocky Mountain Center for Occupational and Environmental Health for study implementation and collaboration: Yuanhang Zhao, Marcus Stucki, Nicole Green, Megan Davidson, Tyler Allison, Naveen Naveed, Melanie Finlay, Christian Guzman, Abby Faha, Charles Schuknecht, Ben Abbey, Becket Harris, Valerie Zwonitzer, Judie Guzman, Soonyoung Kwon, Camil Rowan, Reese Hammer, and Danika Nakai.

Medical writing and editorial support, under the direction of the authors, was provided by Taylor Tibbs, Sara Musetti Jenkins, and John Forbes of RTI Health Solutions and was funded by Novavax, Inc. Editorial support was also provided by Anar Murphy, PhD, CMPP, of Novavax, Inc.

## Author contributions

All authors contributed to the study conception and design. Material preparation and data collection was conducted by SC, DO, RZ, LJ, JC, BP, SY, GE, AP, JM, JG, RC, and MB. RZ, DO, BAP, and JC performed the statistical analyses. All authors collaborated on analysis and interpretation. All authors provided feedback on manuscript drafts and approved the final manuscript. The authors accept accountabilities for all aspects of the work, ensuring questions related to accuracy or integrity are investigated and resolved. All authors had full access to all the data in the study and had final responsibility for the decision to submit for publication.

## Data sharing statement

Study information is available online at: https://www.clinicaltrials.gov/study/NCT06633835. Requests submitted to the corresponding author will be considered upon publication of this article and de-identified participant data might be provided.

## Declaration of interests

SKY has received funding from Novavax, Inc. pertaining to this study and from the Centers for Disease Control and Prevention for prior COVID-19 vaccine studies and has served as an advisory board member, without compensation, for Pfizer-BioNTech.

GLE, AP, JM, JG, and RC are funded through a contract agreement with the University of Utah, with prime funding and study drug provided by Novavax, Inc. SC, DO, RZ, LJ, and JC are full-time employees of RTI Health Solutions, an independent nonprofit research organization, which was retained by Novavax, Inc. to conduct the research. BAP is a former employee of RTI Health Solutions. Their compensation is unconnected to the studies on which they work.

BH, BW, JF, and MDR are Novavax, Inc. employees or contractors and as such receive a work salary and may hold Novavax, Inc. stock.

## Role of the funding source

Funding was provided by Novavax, Inc.

## Notes

### Clinical Trial

NCT06633835

### Clinical Protocols

https://www.clinicaltrials.gov/study/NCT06633835

