## Supplementary Materials for "mRNA and Protein Subunit COVID-19 Vaccine Reactogenicity and the Relationship to Productivity for Healthcare Workers and First Responders"

### SUPPLEMENTAL MATERIAL

#### Table S1. Difference in the percentage of participants experiencing local or systemic reactogenicity symptoms during the first 2 days post vaccination

| | NVX (N=218) | PFZ (N=365) |
|---|---|---|
| **Systemic reactogenicity symptoms** | | |
| Overall | | |
| Percentage, % (95% CI) | 73.6 (65.7 to 81.6) | 87.5 (82.2 to 92.7) |
| Difference in percentages (95% CI) NVX - PFZ | -13.8 (-23.3 to -4.3) | |
| Odds ratio (95% CI) NVX vs PFZ* | 0.401 (0.214 to 0.751) | |
| p value | 0.0044 | |
| Fever[a] | | |
| Percentage, % (95% CI) | 2.3 (-0.5 to 5.1) | 13.8 (8.6 to 19.0) |
| Difference in percentages (95% CI) NVX - PFZ | -11.5 (-17.4 to -5.6) | |
| Odds ratio (95% CI) NVX vs PFZ* | 0.146 (0.039 to 0.555) | |
| p value | 0.0001 | |
| Fatigue[b] | | |
| Percentage, % (95% CI) | 48.9 (39.8 to 58.0) | 68.1 (60.9 to 75.4) |
| Difference in percentages (95% CI) NVX - PFZ | -19.3 (-30.9 to -7.6) | |
| Odds ratio (95% CI) NVX vs PFZ* | 0.447 (0.273 to 0.732) | |
| p value | 0.0012 | |
| Malaise/Feeling Sick[a] | | |
| Percentage, % (95% CI) | 28.9 (20.6 to 37.1) | 49.2 (41.5 to 57.0) |

| | NVX (N=218) | PFZ (N=365) |
|---|---|---|
| Difference in percentages (95% CI) NVX - PFZ | -20.4 (-31.7 to -9.1) | |
| Odds ratio (95% CI) NVX vs PFZ* | 0.418 (0.252 to 0.695) | |
| p value | 0.0004 | |
| Muscle Pain[a] | | |
| Percentage, % (95% CI) | 44.5 (35.3 to 53.6) | 62.9 (55.4 to 70.5) |
| Difference in percentages (95% CI) NVX - PFZ | -18.4 (-30.3 to -6.6) | |
| Odds ratio (95% CI) NVX vs PFZ* | 0.472 (0.289 to 0.772) | |
| p value | 0.0024 | |
| Joint Pain[a] | | |
| Percentage, % (95% CI) | 10.8 (4.5 to 17.1) | 26.4 (19.7 to 33.0) |
| Difference in percentages (95% CI) NVX - PFZ | -15.5 (-24.7 to -6.3) | |
| Odds ratio (95% CI) NVX vs PFZ* | 0.340 (0.163 to 0.710) | |
| p value | 0.001 | |
| Nausea/Vomiting[a] | | |
| Percentage, % (95% CI) | 5.4 (1.3 to 9.6) | 8.1 (3.8 to 12.4) |
| Difference in percentages (95% CI) NVX - PFZ | -2.6 (-8.6 to 3.3) | |
| Odds ratio (95% CI) NVX vs PFZ* | 0.654 (0.244 to 1.756) | |
| p value | 0.3822 | |
| Headache[a] | | |
| Percentage, % (95% CI) | 27.5 (19.2 to 35.7) | 48.6 (40.8 to 56.5) |
| Difference in percentages (95% CI) NVX - PFZ | -21.2 (-32.5 to -9.8) | |
| Odds ratio (95% CI) NVX vs PFZ* | 0.400 (0.238 to 0.671) | |
| p value | 0.0003 | |

| | NVX (N=218) | PFZ (N=365) |
|---|---|---|
| **Local reactogenicity symptoms** | | |
| Overall | | |
| Percentage, % (95% CI) | 82.4 (75.6 to 89.1) | 98.0 (95.7 to 100.2) |
| Difference in percentages (95% CI) NVX - PFZ | -15.6 (-22.7 to -8.5) | |
| Odds ratio (95% CI) NVX vs PFZ* | 0.097 (0.028 to 0.330) | |
| p value | <0.0001 | |
| Injection Site Pain[a] | | |
| Percentage, % (95% CI) | 77.4 (69.9 to 85.0) | 94.8 (91.6 to 98.0) |
| Difference in percentages (95% CI) NVX - PFZ | -17.3 (-25.5 to -9.2) | |
| Odds ratio (95% CI) NVX vs PFZ* | 0.189 (0.087 to 0.412) | |
| p value | <0.0001 | |
| Injection Site Tenderness[a] | | |
| Percentage, % (95% CI) | 74.5 (66.7 to 82.4) | 91.6 (87.3 to 95.9) |
| Difference in percentages (95% CI) NVX - PFZ | -17.1 (-26.0 to -8.1) | |
| Odds ratio (95% CI) NVX vs PFZ* | 0.268 (0.134 to 0.537) | |
| p value | 0.0002 | |
| Injection Site Redness[a] | | |
| Percentage, % (95% CI) | 3.7 (0.2 to 7.3) | 15.2 (9.9 to 20.5) |
| Difference in percentages (95% CI) NVX - PFZ | -11.5 (-17.9 to -5.1) | |
| Odds ratio (95% CI) NVX vs PFZ* | 0.216 (0.074 to 0.630) | |
| p value | 0.0005 | |
| Injection Site Swelling[a] | | |
| Percentage, % (95% CI) | 9.1 (3.8 to 14.5) | 22.1 (15.9 to 28.3) |

| | NVX (N=218) | PFZ (N=365) |
|---|---|---|
| Difference in percentages (95% CI) NVX - PFZ | -13.0 (-21.2 to -4.8) | |
| Odds ratio (95% CI) NVX vs PFZ* | 0.354 (0.169 to 0.740) | |
| p value | 0.0019 | |

CI = confidence interval; NVX = Novavax protein subunit COVID-19 vaccine; PFZ = Pfizer-BioNTech mRNA vaccine.

Note 1: Odds ratio, percentages, CIs, and p value are from a logistic regression for the comparison between the Novavax and Pfizer mRNA groups with covariate adjustments for age, sex at birth, primarily working in a hospital setting, along with the interactions of vaccine group by these covariates.

Note 2: Participants are considered as having experienced no systemic reactogenicity symptoms if they reported grade 0 for all 7 systemic symptoms assessed.

Note 3: Six participants were removed from the analysis sample due to incomplete responses (Novavax: 1 removed due to missing sex; Pfizer: 1 removed for incomplete reactogenicity, 4 removed for missing hospital work setting).

* Reference.

[a] Five participants were removed from the analysis sample due to incomplete responses (Novavax: 1 removed due to missing sex; Pfizer: 4 removed for missing hospital work setting).

[b] Six participants were removed from the analysis sample due to incomplete responses (Novavax: 1 removed due to missing sex; Pfizer: 1 removed for incomplete reactogenicity, 4 removed for missing hospital work setting).

**Table S2. Difference in the mean number of systemic reactogenicity symptoms experienced during the first 2 days post vaccination**

| | NVX (N=218) | PFZ (N=364) |
|---|---|---|
| NVX - PFZ | | |
| Mean number of symptoms (95% CI) | 1.7 (1.5, 2.0) | 2.8 (2.5, 3.1) |
| Difference in the mean number of symptoms (95% CI) NVX – PFZ | -1.0 (-1.4, -0.6) | |
| p value | <0.0001 | |

CI = confidence interval; NVX = Novavax protein subunit COVID-19 vaccine; PFZ = Pfizer-BioNTech mRNA vaccine.

Note 1: Estimates, confidence intervals, and p value are from a negative binomial regression model for the comparison between the Novavax and Pfizer mRNA groups with covariate adjustments for age, sex at birth, primarily working in a hospital setting, along with the interactions of vaccine group by these covariates.

Note 2: The number of systemic reactogenicity symptoms was calculated by summing the number of symptoms each participant reported with a grade 1 or worse.

Note 3: Six participants were removed from the analysis sample due to incomplete responses (Novavax: 1 removed due to missing sex; Pfizer: 1 removed for incomplete reactogenicity, 4 removed for missing hospital work setting).

**Table S3. Difference in the percentage of participants experiencing any grade 2 or worse systemic or local reactogenicity symptoms during the first 2 days post vaccination**

| | NVX (N=218) | PFZ (N=364) |
|---|---|---|
| **Any grade 2 or worse systemic or local reactogenicity symptom[a]** | | |
| NVX - PFZ | | |
| Percentage, % (95% CI) | 31.2 (22.5 to 39.9) | 67.0 (59.4 to 74.6) |
| Difference in percentages (95% CI) NVX - PFZ | -35.8 (-47.4 to -24.2) | |
| Odds ratio (95% CI) NVX vs PFZ* | 0.223 (0.131 to 0.381) | |
| p value | <0.0001 | |
| **Any grade 2 or worse systemic reactogenicity symptom[a]** | | |
| NVX - PFZ mRNA | | |
| Percentage, % (95% CI) | 19.2 (11.8 to 26.6) | 44.5 (36.7 to 52.3) |
| Difference in percentages (95% CI) NVX - PFZ | -25.3 (-36.1 to -14.6) | |
| Odds ratio (95% CI) NVX vs PFZ* | 0.296 (0.167 to 0.524) | |
| p value | <0.0001 | |
| **Any grade 2 or worse local reactogenicity symptom[b]** | | |
| NVX - PFZ mRNA | | |
| Percentage, % (95% CI) | 22.8 (14.8 to 30.8) | 57.7 (49.6 to 65.8) |
| Difference in percentages (95% CI) NVX - PFZ | -34.8 (-46.2 to -23.5) | |
| Odds ratio (95% CI) NVX vs PFZ * | 0.217 (0.124 to 0.381) | |
| p value | <0.0001 | |

CI = confidence interval; NVX = Novavax protein subunit COVID-19 vaccine; PFZ = Pfizer mRNA COVID-19 vaccine.

Note 1: Odds ratio, percentages, confidence intervals, and p value are from a logistic regression for the comparison between the Novavax and Pfizer mRNA groups with covariate adjustments for age, sex at birth, primarily working in a hospital setting, along with the interactions of vaccine group by these covariates.

Note 2: Participants are considered as having experienced any grade 2 or worse reactogenicity symptom if they reported at least one grade 2 or worse for any of the 7 systemic or 4 local symptoms assessed.

*Reference.

[a] Six participants were removed from the analysis sample due to incomplete responses (Novavax: 1 removed due to missing sex; Pfizer: 1 removed for incomplete reactogenicity, 4 removed for missing hospital work setting).

[b] Five participants were removed from the analysis sample due to incomplete responses (Novavax: 1 removed due to missing sex; Pfizer: 4 removed for missing hospital work setting).

**Table S4. Frequency and percentage of participants experiencing individual and overall reactogenicity symptoms during the first 2 days post vaccination cross tabulated during days 3 to 7 post vaccination**

| Symptom | Reactogenicity during days 1 to 2 post vaccination | Reactogenicity during days 3 to 7 post vaccination | NVX (N= 217), n (%) | PFZ (n=366), n (%) |
|---|---|---|---|---|
| Any reactogenicity symptom | No symptoms | No symptoms | 12 (5.5) | 7 (1.9) |
| | | Grade 1 or worse | 2 (0.9) | 0 (0.0) |
| | Grade 1 or worse | No symptoms | 109 (50.2) | 163 (44.5) |
| | | Grade 1 or worse | 94 (43.3) | 195 (53.3) |
| | | Missing | 0 | 1 |
| Any systemic symptom | No symptoms | No symptoms | 48 (22.1) | 43 (11.7) |
| | | Grade 1 or worse | 8 (3.7) | 4 (1.1) |
| | Grade 1 or worse | No symptoms | 90 (41.5) | 168 (45.9) |
| | | Grade 1 or worse | 71 (32.7) | 150 (41.0) |
| | | Missing | 0 | 1 |
| Fever | No symptoms | No symptoms | 204 (94.0) | 313 (85.5) |
| | | Grade 1 or worse | 6 (2.8) | 5 (1.4) |
| | Grade 1 or worse | No symptoms | 7 (3.2) | 44 (12.0) |
| | | Grade 1 or worse | 0 (0.0) | 4 (1.1) |
| Fatigue | No symptoms | No symptoms | 92 (42.4) | 105 (28.7) |

| Symptom | Reactogenicity during days 1 to 2 post vaccination | Reactogenicity during days 3 to 7 post vaccination | NVX (N= 217), n (%) | PFZ (n=366), n (%) |
|---|---|---|---|---|
| | | Grade 1 or worse | 9 (4.1) | 8 (2.2) |
| | Grade 1 or worse | No symptoms | 68 (31.3) | 158 (43.2) |
| | | Grade 1 or worse | 48 (22.1) | 95 (26.0) |
| Malaise/feeling sick | No symptoms | No symptoms | 141 (65.0) | 158 (43.2) |
| | | Grade 1 or worse | 14 (6.5) | 15 (4.1) |
| | Grade 1 or worse | No symptoms | 41 (18.9) | 146 (39.9) |
| | | Grade 1 or worse | 21 (9.7) | 47 (12.8) |
| Muscle pain | No symptoms | No symptoms | 113 (52.1) | 112 (30.6) |
| | | Grade 1 or worse | 10 (4.6) | 12 (3.3) |
| | Grade 1 or worse | No symptoms | 71 (32.7) | 193 (52.7) |
| | | Grade 1 or worse | 23 (10.6) | 48 (13.1) |
| | | Missing | 0 | 1 |
| Joint pain | No symptoms | No symptoms | 181 (83.4) | 264 (72.1) |
| | | Grade 1 or worse | 7 (3.2) | 11 (3.0) |
| | Grade 1 or worse | No symptoms | 18 (8.3) | 76 (20.8) |
| | | Grade 1 or worse | 11 (5.1) | 15 (4.1) |
| Nausea/vomiting | No Symptoms | No Symptoms | 195 (89.9) | 324 (88.5) |

| Symptom | Reactogenicity during days 1 to 2 post vaccination | Reactogenicity during days 3 to 7 post vaccination | NVX (N= 217), n (%) | PFZ (n=366), n (%) |
|---|---|---|---|---|
| | | Grade 1 or Worse | 8 (3.7) | 9 (2.5) |
| | Grade 1 or worse | No symptoms | 10 (4.6) | 27 (7.4) |
| | | Grade 1 or worse | 4 (1.8) | 6 (1.6) |
| Headache | No symptoms | No symptoms | 136 (62.7) | 176 (48.1) |
| | | Grade 1 or worse | 20 (9.2) | 27 (7.4) |
| | Grade 1 or worse | No symptoms | 39 (18.0) | 103 (28.1) |
| | | Grade 1 or worse | 22 (10.1) | 60 (16.4) |
| Any local symptom | No symptoms | No symptoms | 29 (13.4) | 10 (2.7) |
| | | Grade 1 or worse | 4 (1.8) | 1 (0.3) |
| | Grade 1 or worse | No symptoms | 144 (66.4) | 251 (68.6) |
| | | Grade 1 or worse | 40 (18.4) | 104 (28.4) |
| Injection site pain | No symptoms | No symptoms | 39 (18.0) | 17 (4.6) |
| | | Grade 1 or worse | 3 (1.4) | 0 (0.0) |
| | Grade 1 or worse | No symptoms | 145 (66.8) | 269 (73.5) |
| | | Grade 1 or worse | 30 (13.8) | 80 (21.9) |
| Injection site tenderness | No symptoms | No symptoms | 48 (22.1) | 28 (7.7) |
| | | Grade 1 or worse | 2 (0.9) | 2 (0.5) |

| Symptom | Reactogenicity during days 1 to 2 post vaccination | Reactogenicity during days 3 to 7 post vaccination | NVX (N= 217), n (%) | PFZ (n=366), n (%) |
|---|---|---|---|---|
| | Grade 1 or worse | No symptoms | 135 (62.2) | 250 (68.3) |
| | | Grade 1 or worse | 32 (14.7) | 86 (23.5) |
| Injection site redness | No symptoms | No symptoms | 197 (90.8) | 316 (86.3) |
| | | Grade 1 or worse | 7 (3.2) | 4 (1.1) |
| | Grade 1 or worse | No symptoms | 12 (5.5) | 36 (9.8) |
| | | Grade 1 or worse | 1 (0.5) | 10 (2.7) |
| Injection site swelling | No symptoms | No symptoms | 188 (86.6) | 288 (78.7) |
| | | Grade 1 or worse | 5 (2.3) | 5 (1.4) |
| | Grade 1 or worse | No symptoms | 21 (9.7) | 57 (15.6) |
| | | Grade 1 or worse | 3 (1.4) | 16 (4.4) |

NVX = Novavax protein subunit COVID-19 vaccine; PFZ = Pfizer-BioNTech mRNA vaccine.

**Table S5. Disruption of work, social, and family life caused by vaccine symptoms 2 days post vaccination**

| | First 2 days post vaccination[a] | | |
|---|---|---|---|
| | NVX (n=219) | PFZ (n=369) | Overall (n=588) |
| How many hours in the relevant period following your vaccination, did your symptoms cause you to miss work or leave you unable to carry out your normal daily responsibilities? | | | |
| Mean (SD) | 0.7 (2.98) | 1.4 (4.85) | 1.1 (4.27) |
| How many hours in the relevant period following your vaccination did you feel so impaired by your symptoms, that even though you went to work, your productivity was reduced? | | | |
| Mean (SD) | 0.8 (3.07) | 2.4 (5.09) | 1.8 (4.51) |
| The vaccine symptoms have disrupted work/schoolwork, n (%) | | | |
| Not at all (0) | 119 (54.3) | 145 (39.3) | 264 (44.9) |
| Mildly (1) | 21 (9.6) | 40 (10.8) | 61 (10.4) |
| Mildly (2) | 8 (3.7) | 38 (10.3) | 46 (7.8) |
| Mildly (3) | 10 (4.6) | 27 (7.3) | 37 (6.3) |
| Moderately (4) | 2 (0.9) | 18 (4.9) | 20 (3.4) |
| Moderately (5) | 9 (4.1) | 12 (3.3) | 21 (3.6) |
| Moderately (6) | 0 (0.0) | 9 (2.4) | 9 (1.5) |
| Markedly (7) | 2 (0.9) | 6 (1.6) | 8 (1.4) |
| Markedly (8) | 1 (0.5) | 4 (1.1) | 5 (0.9) |
| Markedly (9) | 0 (0.0) | 0 (0.0) | 0 (0.0) |
| Extremely (10) | 0 (0.0) | 4 (1.1) | 4 (0.7) |
| Did no work/schoolwork during the period for reasons unrelated to the vaccine | 47 (21.5) | 66 (17.9) | 113 (19.2) |
| The vaccine symptoms have disrupted your social life / leisure activities, n (%) | | | |
| Not at all (0) | 159 (72.6) | 212 (57.5) | 371 (63.1) |

| | First 2 days post vaccination[a] | | |
|---|---|---|---|
| | NVX (n=219) | PFZ (n=369) | Overall (n=588) |
| Mildly (1) | 28 (12.8) | 34 (9.2) | 62 (10.5) |
| Mildly (2) | 9 (4.1) | 31 (8.4) | 40 (6.8) |
| Mildly (3) | 11 (5.0) | 33 (8.9) | 44 (7.5) |
| Moderately (4) | 3 (1.4) | 15 (4.1) | 18 (3.1) |
| Moderately (5) | 4 (1.8) | 19 (5.1) | 23 (3.9) |
| Moderately (6) | 2 (0.9) | 7 (1.9) | 9 (1.5) |
| Markedly (7) | 2 (0.9) | 11 (3.0) | 13 (2.2) |
| Markedly (8) | 1 (0.5) | 3 (0.8) | 4 (0.7) |
| Markedly (9) | 0 (0.0) | 1 (0.3) | 1 (0.2) |
| Extremely (10) | 0 (0.0) | 3 (0.8) | 3 (0.5) |
| The vaccine symptoms have disrupted your family life / home responsibilities, n (%) | | | |
| Not at all (0) | 156 (71.2) | 189 (51.2) | 345 (58.7) |
| Mildly (1) | 25 (11.4) | 52 (14.1) | 77 (13.1) |
| Mildly (2) | 15 (6.8) | 37 (10.0) | 52 (8.8) |
| Mildly (3) | 8 (3.7) | 33 (8.9) | 41 (7.0) |
| Moderately (4) | 6 (2.7) | 12 (3.3) | 18 (3.1) |
| Moderately (5) | 1 (0.5) | 11 (3.0) | 12 (2.0) |
| Moderately (6) | 3 (1.4) | 12 (3.3) | 15 (2.6) |
| Markedly (7) | 4 (1.8) | 13 (3.5) | 17 (2.9) |
| Markedly (8) | 1 (0.5) | 7 (1.9) | 8 (1.4) |
| Markedly (9) | 0 (0.0) | 1 (0.3) | 1 (0.2) |
| Extremely (10) | 0 (0.0) | 2 (0.5) | 2 (0.3) |
| Total score of global impairment (Sheehan disability score) | | | |
| Mean (SD) | 2.2 (4.40) | 4.6 (6.32) | 3.7 (5.80) |
| Median (IQR) | 0.0 (3.0) | 2.0 (7.0) | 0.0 (6.0) |

IQR = interquartile range; NVX=Novavax protein subunit COVID-19 vaccine; PFZ=Pfizer-BioNTech mRNA vaccine; SD = standard deviation.

Note: The total score of global impairment ranges from 0–30, with higher scores indicating greater impairment.

[a] The results are derived from questions 12 to 16 among participants who completed the Day 2 post-vaccination questionnaire.

**Table S6. Difference in the mean total score of global impairment of work, social, and family life disruption caused by vaccine symptoms during the first 2 days post vaccination (Sheehan Disability Scale score)**

| | NVX (N=218) | PFZ (N=365) |
|---|---|---|
| NVX - PFZ | | |
| Least-squares (LS) mean (95% CI) | 1.9 (0.9 to 2.9) | 4.5 (3.7 to 5.4) |
| Difference in LS means (95% CI) Novavax – Pfizer mRNA | -2.6 (-3.9 to -1.3) | |
| p value | 0.0001 | |

CI = confidence interval; LS = least squares; NVX = Novavax protein subunit COVID-19 vaccine; PFZ = Pfizer-BioNTech mRNA vaccine.

Note 1: Estimates, confidence intervals, and p value are from an ANOVA model for the comparison between the Novavax and Pfizer mRNA groups with covariate adjustments for age, sex at birth, primarily working in a hospital setting, along with the interactions of vaccine group by these covariates.

Note 2: The total score of global impairment ranges from 0–30, with higher scores indicating greater impairment.

Note 3: Five participants were removed from the analysis sample due to incomplete responses (Novavax: 1 removed due to missing sex; Pfizer: 4 removed for missing hospital work setting).

**Table S7. Disruption of work, social, and family life caused by vaccine symptoms days 3–7 post vaccination**

| | Days 3-7 Post vaccination[a] | | |
|---|---|---|---|
| | **NVX (n=217)** | **PFZ (n=366)** | **Overall (n=583)** |
| How many hours in the relevant period following your vaccination, did your symptoms cause you to miss work or leave you unable to carry out your normal daily responsibilities? | | | |
| Mean (SD) | 1.1 (6.64) | 0.8 (4.36) | 0.9 (5.32) |
| Median (IQR) | 0.0 (0.0) | 0.0 (0.0) | 0.0 (0.0) |
| Min, max | 0, 73 | 0, 36 | 0, 73 |
| How many hours in the relevant period following your vaccination did you feel so impaired by your symptoms, that even though you went to work, your productivity was reduced? | | | |
| Mean (SD) | 1.1 (6.26) | 1.0 (4.54) | 1.0 (5.24) |
| Median (IQR) | 0.0 (0.0) | 0.0 (0.0) | 0.0 (0.0) |
| Min, max | 0, 72 | 0, 48 | 0, 72 |
| The vaccine symptoms have disrupted work/schoolwork, n (%) | | | |
| Not at all (0) | 174 (80.2) | 300 (82.0) | 474 (81.3) |
| Mildly (1) | 17 (7.8) | 22 (6.0) | 39 (6.7) |
| Mildly (2) | 7 (3.2) | 13 (3.6) | 20 (3.4) |
| Mildly (3) | 5 (2.3) | 6 (1.6) | 11 (1.9) |
| Moderately (4) | 4 (1.8) | 5 (1.4) | 9 (1.5) |
| Moderately (5) | 0 (0.0) | 3 (0.8) | 3 (0.5) |
| Moderately (6) | 2 (0.9) | 1 (0.3) | 3 (0.5) |
| Markedly (7) | 2 (0.9) | 3 (0.8) | 5 (0.9) |
| Markedly (8) | 1 (0.5) | 1 (0.3) | 2 (0.3) |
| Markedly (9) | 0 (0.0) | 0 (0.0) | 0 (0.0) |
| Extremely (10) | 0 (0.0) | 1 (0.3) | 1 (0.2) |

| | Days 3-7 Post vaccination[a] | | |
|---|---|---|---|
| | NVX (n=217) | PFZ (n=366) | Overall (n=583) |
| Did no work/school work during the period for reasons unrelated to the vaccine | 5 (2.3) | 11 (3.0) | 16 (2.7) |
| The vaccine symptoms have disrupted your social life / leisure activities, n (%) | | | |
| Not at all (0) | 181 (83.4) | 311 (85.0) | 492 (84.4) |
| Mildly (1) | 10 (4.6) | 19 (5.2) | 29 (5.0) |
| Mildly (2) | 13 (6.0) | 13 (3.6) | 26 (4.5) |
| Mildly (3) | 6 (2.8) | 9 (2.5) | 15 (2.6) |
| Moderately (4) | 0 (0.0) | 3 (0.8) | 3 (0.5) |
| Moderately (5) | 2 (0.9) | 4 (1.1) | 6 (1.0) |
| Moderately (6) | 3 (1.4) | 3 (0.8) | 6 (1.0) |
| Markedly (7) | 2 (0.9) | 3 (0.8) | 5 (0.9) |
| Markedly (8) | 0 (0.0) | 1 (0.3) | 1 (0.2) |
| Markedly (9) | 0 (0.0) | 0 (0.0) | 0 (0.0) |
| Extremely (10) | 0 (0.0) | 0 (0.0) | 0 (0.0) |
| The vaccine symptoms have disrupted your family life / home responsibilities, n (%) | | | |
| Not at all (0) | 188 (86.6) | 311 (85.0) | 499 (85.6) |
| Mildly (1) | 8 (3.7) | 21 (5.7) | 29 (5.0) |
| Mildly (2) | 6 (2.8) | 12 (3.3) | 18 (3.1) |
| Mildly (3) | 5 (2.3) | 8 (2.2) | 13 (2.2) |
| Moderately (4) | 2 (0.9) | 2 (0.5) | 4 (0.7) |
| Moderately (5) | 2 (0.9) | 5 (1.4) | 7 (1.2) |
| Moderately (6) | 3 (1.4) | 3 (0.8) | 6 (1.0) |
| Markedly (7) | 2 (0.9) | 2 (0.5) | 4 (0.7) |
| Markedly (8) | 1 (0.5) | 1 (0.3) | 2 (0.3) |
| Markedly (9) | 0 (0.0) | 1 (0.3) | 1 (0.2) |
| Extremely (10) | 0 (0.0) | 0 (0.0) | 0 (0.0) |
| Total score of global impairment (Sheehan Disability Scale score) | | | |
| Mean (SD) | 1.3 (3.77) | 1.2 (3.66) | 1.3 (3.70) |

| | Days 3-7 Post vaccination[a] | | |
|---|---|---|---|
| | **NVX (n=217)** | **PFZ (n=366)** | **Overall (n=583)** |
| Median (IQR) | 0.0 (0.0) | 0.0 (0.0) | 0.0 (0.0) |

IQR = interquartile range; NVX=Novavax protein subunit COVID-19 vaccine; PFZ=Pfizer-BioNTech mRNA vaccine; SD = standard deviation.

Note: The total score of global impairment ranges from 0–30, with higher scores indicating greater impairment.

[a] The results are derived from questions 45 to 49 among participants who completed the first 2 days post vaccination and the Days 3–7 post-vaccination questionnaires.

#### Figure S1. Study Design

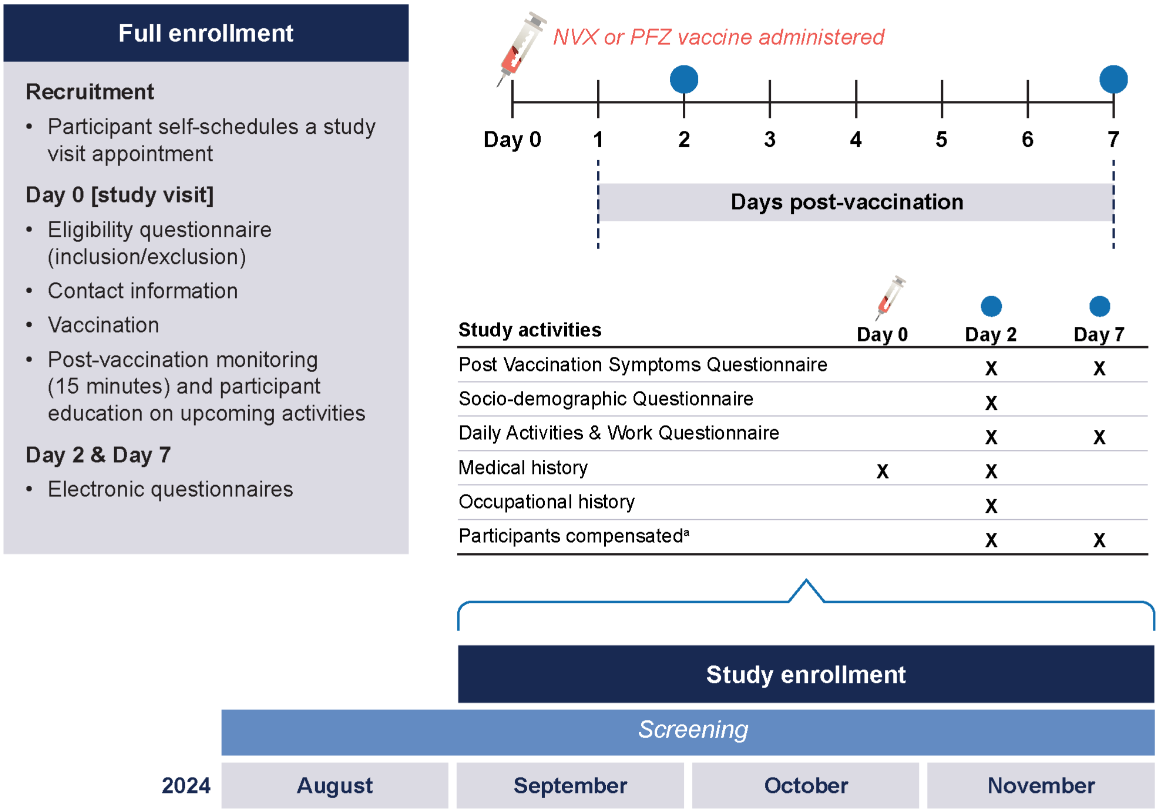

| Study activities | Day 0 | Day 2 | Day 7 |
|---|---|---|---|
| Post Vaccination Symptoms Questionnaire | | X | X |
| Socio-demographic Questionnaire | | X | |
| Daily Activities & Work Questionnaire | | X | X |
| Medical history | X | X | |
| Occupational history | | X | |
| Participants compensated[a] | | X | X |

NVX=Novavax protein subunit COVID-19 vaccine; PFZ=Pfizer mRNA COVID-19 vaccine.

[a] Participants also compensated at study end
